# Understanding the Experiences and Needs of LGBTIQA+ Individuals when Accessing Abortion Care and Pregnancy Options Counselling: A scoping review

**DOI:** 10.1101/2022.09.06.22279629

**Authors:** Sally Bowler, Kari Vallury, Ernesta Sofija

## Abstract

**Background:** Safe, accessible, and inclusive abortion care and pregnancy options counselling are essential components of sexual and reproductive health and rights. Recent research has documented LGBTIQA+ people are as or more likely than the general population of women to experience an abortion in their lifetime yet face significant barriers to accessing abortion and related care which undermine wellbeing.

**Aims:** The present study undertakes a scoping review of research on the needs and experiences of the LGBTIQA+ population when accessing abortion care, pregnancy options, and post-abortion counselling, to support improved understanding of pregnant people’s preferences, needs, and experiences.

**Materials and Methods:** Online academic databases were searched using terms relating to gender identity and sexuality, abortion, pregnancy-options, and post-abortion counselling to identify peer reviewed papers published in English, from which we selected six publications from the United States, and one from Colombia that described experiences of LGBTIQA+ people accessing abortion-related care.

**Results:** Four of the seven studies reported in-depth or semi-structured interview studies while the remaining three examined cross-sectional surveys. Thematic analysis of all studies highlighted frequent discrimination and exclusion experienced by participants, healthcare avoidance, unsafe abortion, non-disclosure to providers, provision of poor quality of care, and poor health outcomes for LGBTIQA+ people.

**Conclusions:** Gender-inclusive services and training for health providers are key to the provision of safe and accessible abortion care, and imperative to overcome generations of mistrust held by the LGBTIQA+ community. Research into the needs of LGBTIQA+ people when accessing pregnancy options counselling is critically needed.

**Key messages:**

- LGBTIQA+ people experience exclusion, isolation, misgendering, and denial of care when accessing abortion, and barriers faced are compounded by socioeconomic status and race.
- Poor provision of abortion care and negative experiences for this population result in deep mistrust of providers, non-disclosure of gender/sexuality, healthcare avoidance, and self-induced abortion.
- Appropriate, inclusive, and accessible care requires gender affirming and inclusive services, educational materials, and intake forms, community consultation, and improved healthcare provider training and knowledge.

## Introduction

Providing access to safe, inclusive, and comprehensive abortion care is fundamental to the realisation of sexual and reproductive health and rights (SRHR) (Amnesty International, 2021; World Health Organization [WHO], 2021). Safe abortion care and pregnancy-options counselling are vital components of reproductive health care (WHO, 2021). Despite widespread decriminalisation in Australia and around the world, accessibility remains difficult in many places and especially for marginalised population groups.

Inaccessibility of abortion care has direct links to mortality rates due to unsafe abortion (Amnesty International, 2021). According to the WHO (2021), globally, three out of 10 pregnancies end in abortion and almost half of those are unsafe. In higher-income countries, lack of access to quality abortion care is driven by availability of quality health services, abortion legislation, geographical access, financial affordability, confidentiality concerns, misinformation, stigma, and structural barriers (Assifi et al., 2020b; Dawson at al., 2016). This can result in preventable mortality and morbidity, and a suite of health and social consequences for people who cannot safely access abortion services (Dawson et al., 2016; Gerdts et al., 2015; Miller, Wherry & Foster 2020).

Abortion care has historically been described as a service sought by and provided for heterosexual, cisgender women, an assumption that remains common to this day (Amnesty International, 2021). However, studies show high rates of demand for abortion care among LGBTIQA+ people, with access issues compounded by stigma, discrimination, abuse, prejudice, socioeconomic status, and race (Agénor et al., 2021; Charlton et al., 2020; LGBTIQ+ Health Australia, 2021; Lindley & Walsemann, 2015; Saewyc, 2014). It is estimated that 24% of women will have an abortion by age 45, and between one quarter and one third of Australian women will experience at least one abortion in their lifetime (Children by Choice, 2021; Government of South Australia, 2020; Jones & Jerman, 2017; Keogh, Gurrin & Moore, 2021). Studies have found sexual minority women and transgender men are as or more likely to experience both unplanned pregnancies and abortions in comparison to other women, likely due to barriers when accessing sexual and reproductive health services and contraception, and rates of sexual violence (Assifi et al., 2020a; Everett, McCabe & Hughes, 2017; Charlton et al., 2020; Jones, Jerman & Charlton, 2018; Pallito et al., 2013). LGBTIQA+ people are significantly more likely to experience sexual assault, rape, stalking, and/or physical violence in their lifetime than heterosexual women (Australian Human Rights Commission, 2014; Flores et al., 2020; Saewyc et al., 1999).

Despite an estimated 3.2% of Australian adults identifying as bisexual or homosexual, and 2.4% identifying as a different sexual orientation or gender identity (AIHW, 2018) research has found that sexual and reproductive health services are often still unsafe and non-inclusive for this community. Lack of health provider knowledge, gender-related discrimination in health clinics, and refusal of care for people who present as transgender, nonbinary, and/or gender expansive undermines accessibility and quality of care (Moseson et al., 2021c).

The LGBTIQA+ population is under-recognised in guidelines surrounding provision of abortion-related care (Dawson & Leong, 2020), and it is unclear what literature is available that highlights the specific experiences and needs of this population when accessing abortion-related services. This includes services to assist in decision making, abortion-related procedures, and pre- and post-abortion care. For these reasons, a scoping review was conducted to highlight research done in this area, collate best practices in the provision of appropriate, accessible, safe, and inclusive care, and to identify any existing gaps in knowledge. The following research question was formulated: What is known about the experiences and needs of LGBTIQA+ people when accessing abortion care, pregnancy options and/or post-abortion counselling?

While sexual and gender diverse individuals are represented by a range of terms and acronyms, we have elected to use LGBTIQA+ throughout this paper, which was endorsed by the Australian Institute of Family Studies (AIFS) (2021) in consultation with prominent LGBTIQA+ community-led organisation Queerspace (AIFS, 2021), and stands for lesbian, gay, bisexual, transgender, gender diverse, intersex, queer, asexual, and questioning.

## Methods

The reporting of this scoping review is guided by the Preferred Reporting Items for Systematic Reviews and Meta-Analyses extension of Scoping Reviews (PRISMA-ScR) (Tricco et al., 2018).

### Inclusion and exclusion criteria

Studies of all research designs and methodologies including quantitative, qualitative, and mixed method studies were eligible for inclusion to consider the different methods of measuring experiences and needs of abortion related care/services. Peer-reviewed journal papers were included if they were: written in English, involved participants who identified as lesbian, gay, bisexual, transgender, queer, intersex, asexual, and any other person who identifies as a gender or sexuality other than cisgender male, cisgender female, and heterosexual, and described experiences with or preferences regarding pregnancy options or post abortion counselling, or abortion-related care/services. Papers were excluded if they did not fit the population (people other than LGBTIQA+), non-primary research, non-English language studies due to the capacity of the research team, those not pertaining to abortion services or pregnancy options counselling, and not measuring experiences or needs (for example, prevalence studies were not included).

### Search strategy

Seven electronic databases were searched in April 2022, including CINAHL, PUBMed, ProQuest Central, PsychInfo, ClinicalKey, ScienceDirect and Scopus, with support from a specialist research librarian (Max Evett) and further refined in discussions with the research team. The final search strategy for PUBMed can be seen in Table 1. Reference lists of relevant papers were also hand-searched for additional papers, along with searching the Children by Choice website. The search results from each database were exported into EndNote 20 and duplicates were removed.

**Table 1.**
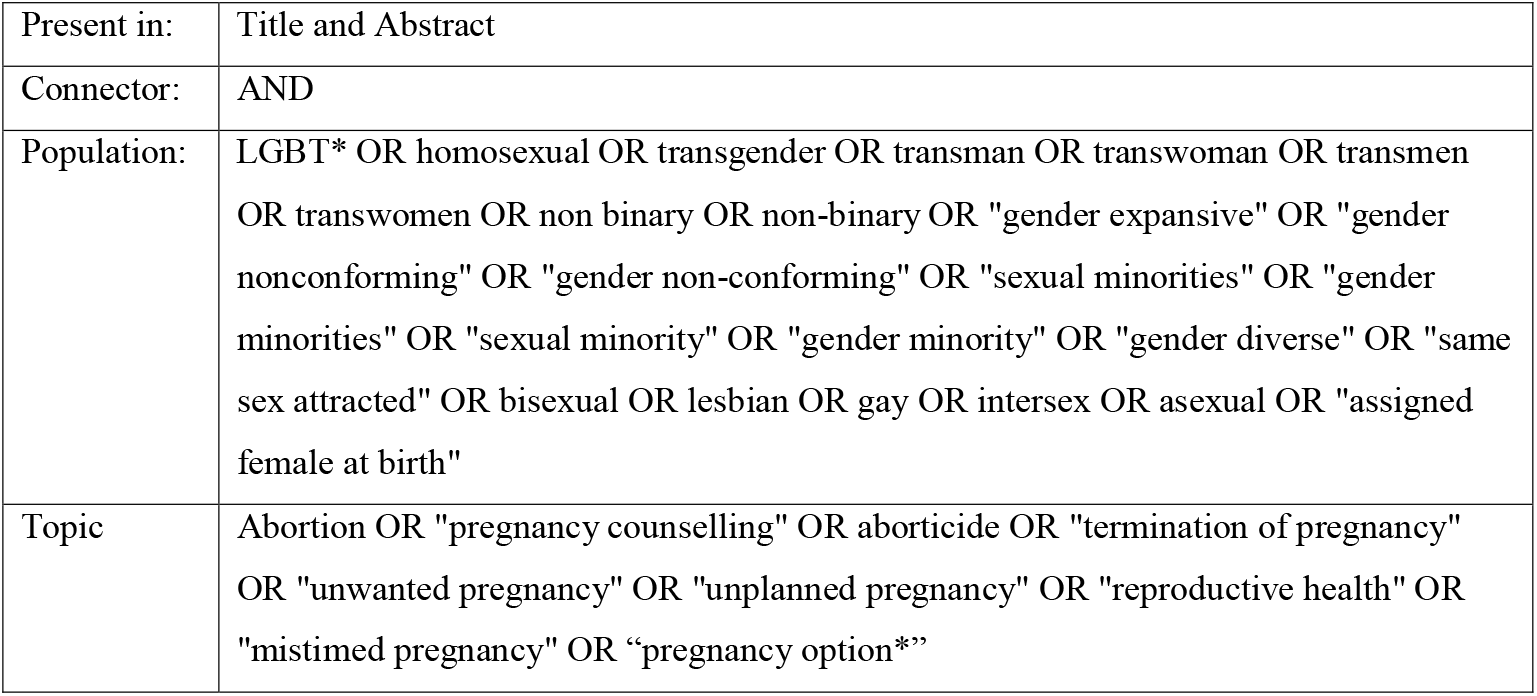
PUBMed Search Strategy (Literature search performed: April 2022)

### Study selection and data synthesis

Citations were screened against inclusion and exclusion criteria and titles and abstracts were assessed for relevance by two reviewers (KV, SB) with indisputably irrelevant studies removed at this stage. The full texts of remaining citations were assessed against the inclusion and exclusion criteria and any disagreements between reviewers were discussed and resolved. Data extraction was then completed using a jointly developed template by the two reviewers to determine variables to extract. Each reviewer independently extracted information regarding the study citation, methodology, participants, location, aims, primary outcome measures and key findings, to continuously discuss and compare the data-charting. A thematic analysis technique was used to synthetize the data into emerging themes, summarizing broad findings.

## Results

### Study selection

Database and hand-searches returned 2550 studies. 2130 records were screened by title and abstract with 47 studies retained for full text review. Forty studies were excluded during full text review (for reasons see Figure 1), and seven studies were included in the final review (See Figure 1) (Carpenter, 2021; Calderón-Jaramillo et al., 2020; Fix et al., 2020; Moseson et al., 2021a; Moseson et al., 2021b; Porsch et al., 2016; Wingo et al., 2018).

**Figure 1.**
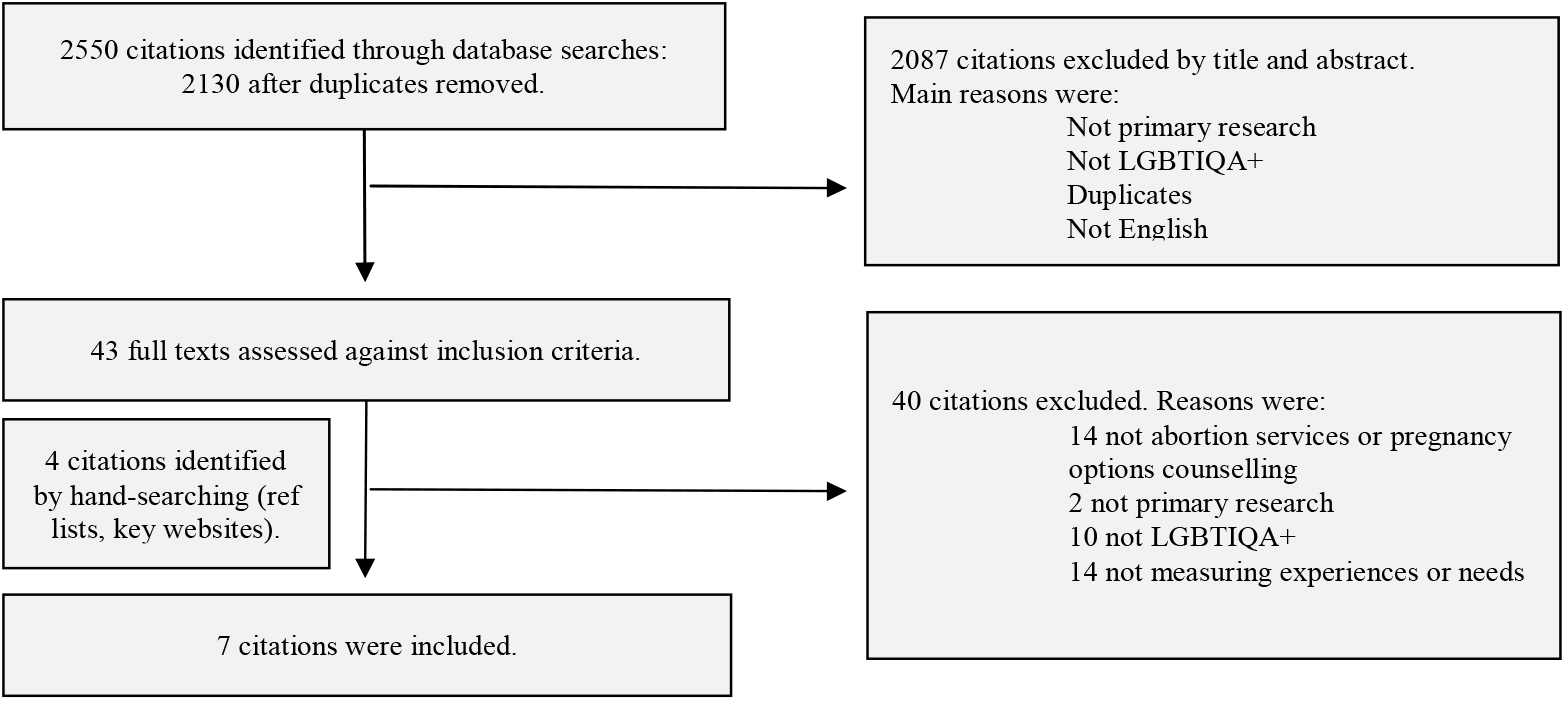
PRISMA flowchart

### Study Characteristics

Of the seven included studies, three conducted qualitative semi-structured in-depth interviews, one conducted focus groups and semi-structed interviews, one used a quantitative cross-sectional survey, and two studies used cross-sectional surveys with quantitative as well as open ended questions. Six studies were conducted in the United States and one in Colombia. All studies included participants who fell under the LGBTIQA+ umbrella. One study also included participants who were relevant stakeholders such as clinicians, advocates, educators, and researchers. Study characteristics and key findings of each paper are recorded in Table 2.

**Table 2.**
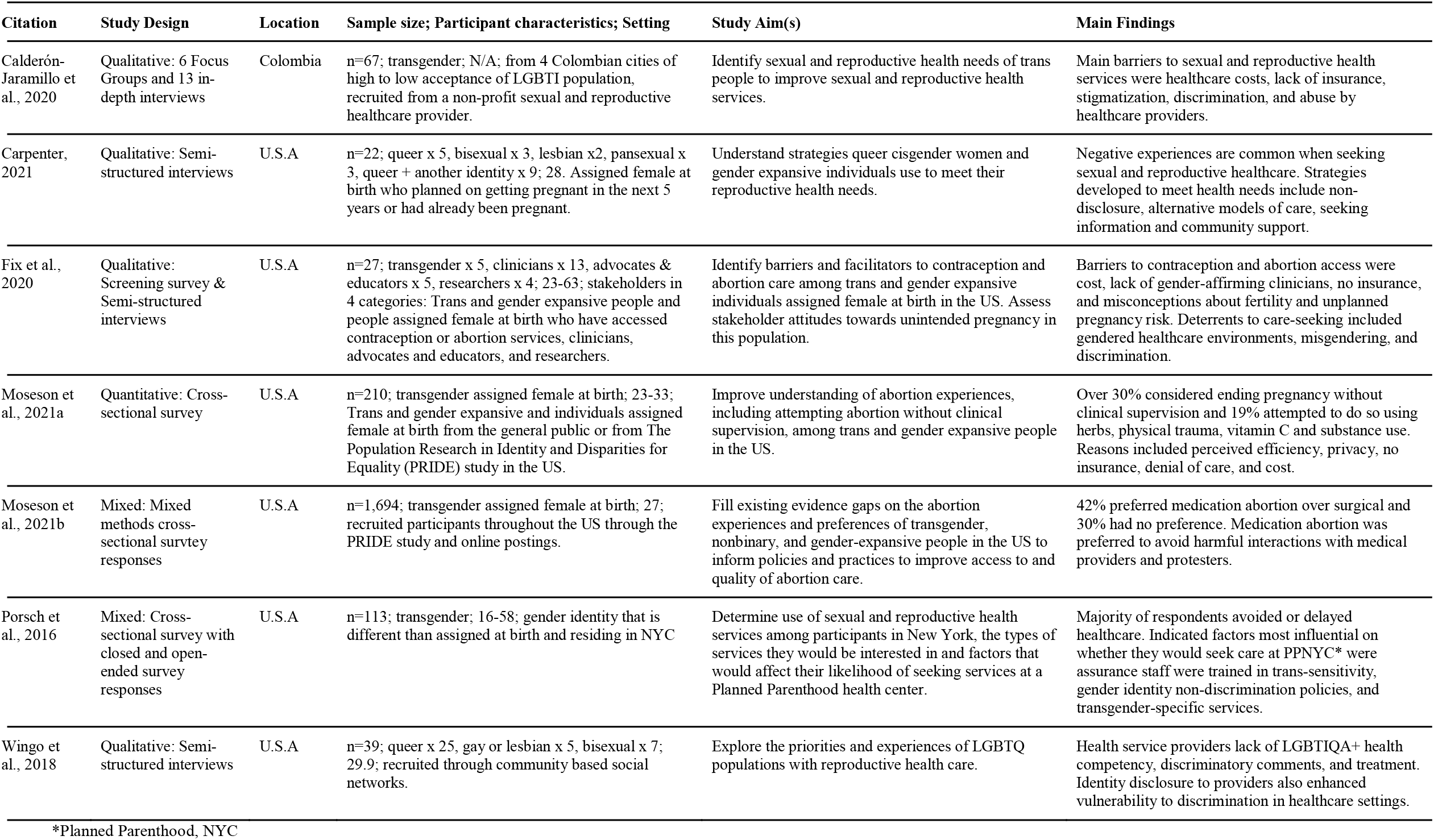
Characteristics of Included Studies.

### Summary of Results

Extracted findings were inductively coded with four overarching themes emerging: 1) experiences when seeking abortion care, 2) identifying inclusive abortion care and information, 3) impacts of poor care, and 4) provision of appropriate, inclusive, and accessible care. There was a paucity of references to pregnancy options and post-abortion counselling, the implications of which are discussed below.

### Experiences seeking abortion care

Participants across included studies reported shared experiences of exclusion, harm, and resilience. Negative opinions about trans people were described as having become essentialised or ‘normalized’ at a societal level and as such embedded in health care seeking experiences (Calderón-Jaramillo et al. 2020). For others, LGBTIQA+ discrimination was seen to be compounded by intersecting oppressions relating to other aspects of their identities. For example, black participants identified needing to consider both their race and ethnicity as well as their gender identity when seeking and choosing abortion care (Fix et al., 2020).

Participants in three studies reported feeling isolated and invisible due to vague or imprecise language used by health providers and a lack of available information adapted for their specific needs (Carpenter, 2021; Fix et al., 2020; Wingo et al., 2018). Ongoing self-advocacy regarding pro-nouns and identity, followed by continued lack of recognition, led to feelings of disempowerment and helplessness among some participants (Fix et al., 2020). Reflective of structural and systemic factors, providers were evidently inadequately trained in the delivery of gender-affirming and inclusive care and frequently described by participants as inexperienced in the sensitive use of de-gendered language and preferred pronouns (Fix et al., 2020).

Along with inappropriate language use, participant in most studies described experiencing discomfort during interactions with providers due to biases or pathologizing attitudes, pressure to fit into sexual binaries, and abuse targeted at their sexual or gender diversity (Calderón-Jaramillo et al., 2020; Carpenter, 2021; Fix et al., 2020; Moseson et al., 2021a; Porsch et al., 2016). Negative experiences impacted future disclosure of sexual and gender identities, which would have otherwise been important to the care they were seeking (Carptenter 2021; Wingo et al., 2018).

Health system organisation provided additional barriers to safe care for participants. Accessing sexual and reproductive health care that was frequently situated in “Women’s health clinics” resulted in anxiety and feelings of being out of place (Fix et al., 2020; Wingo et al., 2018). Additionally, transgender individuals reported having to pay out of pocket for some services because ‘male’ as a gender marker affected certain elements of their health insurance coverage (Fix et al., 2020). Systemic exclusion was further reflected in outdated protocols and limited gender binaries on intake forms (Calderón-Jaramillo et al., 2020; Wingo et al., 2018).

### Identifying inclusive abortion care and information

Participants used a range of strategies to navigate and identify inclusive and safe abortion care and information to support their self-advocacy and fulfill their health needs, sometimes outside of the healthcare system (Carpenter, 2021). Word of mouth and resource sharing were reported as ways of locating queer-identified or queer-friendly providers (Carpenter, 2021; Fix et al., 2020). Some participants sought queer-informed homebirth midwives and doulas for abortion care and pregnancy. One participant, for example, described a “shared understanding” between Queer clients and providers, whereby “unspoken norms and assumptions…allow you to get better care” (Carpenter, 2021). This was further highlighted by Porsch et al. (2016), where 35% of participants said they accessed their care needs at “LGBT health clinics”.

Participants experiences regarding the availability and appropriateness of online resources varied. A lack of online LGBTIQA+ specific resources was interpreted as symbolic of a lack of inclusion: “*when the internet doesn’t have answers for you, that’s when you know you are in some deep territory”* (Carpenter, 2021, p. 480). Overall, a lack of formal information pertaining to inclusive abortion services forced participants to seek information from the LGBTIQA+ community and providers operating outside of formal abortion services.

### Impacts of Poor Care

The impacts of poor quality abortion care described in the included studies were varied and significant. The frequency at which members of the LGBTIQA+ community experienced poor care was seen to have resulted in a “deep mistrust” of health providers, which was exacerbated during each healthcare encounter during which they feel misunderstood and disrespected (Calderón-Jaramillo et al., 2020). Participants reported carefully weighing up the perceived benefits and risks of disclosure in each interaction with healthcare services, with non-disclosure a common strategy for avoiding stigmatisation and discrimination (Fix et al., 2020; Carpenter, 2021). Non-disclosure was seen to have short-term benefits, including avoiding transphobia and stigma, along with long-term negative impacts such as a lack of appropriately tailored medical care (Carpenter 2021, Fix et al., 2020). In some instances, non-disclosure of gender identity meant allowing providers to make assumptions or explicitly hiding sexual orientation or gender identity (Carpenter 2021).

Avoiding healthcare settings out of fear of prejudice and judgment was a common response for those who felt negative experiences would further impact their overall health (Fix et al., 2020). In one study, 74% of participants avoided healthcare in the previous year (Porsch et al., 2016). The reasons for avoiding care included negative experiences with healthcare in the past (68%), lack of healthcare provider knowledge for their needs (65%), a lack of transgender sensitivity (48%), not knowing where to get services (32%), non-inclusive insurance for specific needs (44%) and affordability (45%) (Porsch et al., 2016). For some, this avoidance led to self-induction of abortion without appropriate healthcare and support (Fix et al. 2020; Moseson et al., 2021a). One study found that more than one in three participants considered ending an abortion themselves and nearly one in five attempted a self-managed abortion through physical trauma, ingesting herbs, testosterone use, and other unsafe methods (Moseson et al., 2021a). Moseson and colleagues (2021b) reported 42% of participants who had an abortion preferred medication abortion, largely to avoid further traumatization and discrimination experienced from healthcare provider interactions (Moseson et al., 2021b).

### Provision of Appropriate, Inclusive, and Accessible Care

Best practices for inclusive, safe, and accessible abortion provision at both systemic and provider levels were identified. Potential adaptations to healthcare systems to improve the quality of care were described as including gender-affirming educational materials for individuals, the adoption of gender inclusive intake forms, consistently seeking feedback from the community, the ability for privacy and discretion when coming and going from clinics, better geographic accessibility, and advocacy for comprehensive insurance and contraception (Calderón-Jaramillo et al., 2020; Fix et al., 2020; Moseson et al., 2021b; Wingo et al., 2018).

Knowing their healthcare providers had received training in LGBTIQA+ specific needs and were able to sensitively serve individuals with equality, respect, empathy, and dignity, and with knowledge on the diversity of transitions, was particularly important to research participants and strongly influenced their decisions to access care at such services (Calderón-Jaramillo et al., 2020; Porsch et al., 2016). Alternately, a lack of training of providers drove participants to question the accuracy of medical advice they receive (Calderón-Jaramillo et al., 2020). Studies identified a clear need and demand for queer-friendly and/or queer identified service providers (Carpenter, 2021).

Gender affirming and inclusive language was seen to be crucial to inclusive care provision. For example, asking about a client’s preferred language when referencing body parts, such as “opening” instead of vagina, can be particularly important to transgender people (Fix et al., 2020; Moseson et al., 2021a; Moseson et al., 2021b; Porsch et al., 2016). Gender inclusive, as opposed to gender neutral language, was seen to be more affirming to individual identity (Fix et al., 2020).

### Pregnancy Options and Post-abortion Counselling

Pregnancy options and post-abortion counselling for the LGBTIQA+ population was largely unaddressed in the included studies and was only briefly referred to in one study (Fix et al., 2020). Participants described the likelihood of healthcare providers inappropriately counselling transgender expansive individuals on unintended pregnancy due to poor knowledge and uncompassionate care (Fix et al., 2020). One participant of the same study described they felt supported by their abortion provider and that they saved their life through post-abortion counselling that also addressed their experience of sexual assault (Fix et al., 2020).

## Discussion

This review set out to identify the existing literature on the needs and experiences of the LGBTIQA+ community when accessing pregnancy options counselling and abortion services. Seven relevant studies were identified, six of which were from the USA, indicating a potential lack of generalisability to other countries and their contexts. While disparities in health outcomes between LGBTIQA+ and cis-gender, heterosexual populations are likely universal, barriers to access such as laws, policies, stigma, costs, and insurance vary greatly within and between countries. As such the review highlights significant gaps in research, which requires attention.

Our findings on LGBTIQA+ experiences of abortion care align with the broader literature, which describes the community’s frequent exposure to violence, homophobia, exclusion, anticipated stigma, and emotional harm in health and help-seeking interactions (Agénor et al., 2020; Perales, 2016). These experiences contribute to gender and sexuality non-disclosure by LGBTIQA+ people in healthcare settings (Australian Human Rights Commission, 2014), which implicates their wellbeing. This review found a lack of inclusive information and service provision is driving this population to seek support outside of mainstream health systems and increasing the likelihood of unsafe self-induced abortion. There are undeniably myriad factors that must be tackled at the social, structural, organisational, and individual levels to realise the sexual and reproductive rights of all people (Bateson, Black & Sawleshwarkar, 2019; Charlton et al., 2020). For LGBTIQA+ people, disadvantage and discrimination is often compounded by other marginalised identities such as race, class, age, ethnicity, religion, location, and disability. This impacts the accessibility of both abortion and sexual and reproductive health services (Logie et al., 2018; Moegelin et al., 2010), resulting in poorer health outcomes for LGBTIQA+ people, including increased mental health concerns and healthcare avoidance (Agénor et al., 2020; Calderón-Jaramillo et al., 2020; Carptenter, 2021; Charlton et al., 2020). Specifically, cost has been shown to be a particular barrier to health care access for LGBITQA+ people in America (Porsch et al., 2016), who face lower rates of employment than the general population, and subsequently have reduced access to health insurance and finances to support abortion access (Rosentel, VandeVusse & Hill, 2020). Structural interventions, such as the revision of health insurance regulations and advocacy to support the financing of services for people of all genders and sexualities, are thus critical (Calderón-Jaramillo et al., 2020; Wingo et al., 2018).

### Implications for policy and practice

The included studies identified a range of strategies which are likely to have significant impacts on the health outcomes for LGBTIQA+ people with minimal resourcing required. During interactions between structural and individual factors, tactics can be implemented to prevent continued harm. For example, within organisations, the use of binary gender language in communication materials reinforces social discrimination and compounds feelings of isolation and invisibility for LGBTIQA+ people (Logie et al., 2018). Training new healthcare providers within discriminatory social and organizational settings leads to the continued provision of poor-quality care (Fix et al., 2020). As such, strategies to improve the safety and accessibility of care must be multi-level and co-designed.

The development of gender-inclusive health services to signal safety to this community include improving knowledgeability of providers, adopting inclusive language within organisations both internally and in external communications, and offering an array of services to meet diverse needs. With only 23% of all U.S. clinics offering transgender-specific health services (Jones et al., 2020), transgender abortion seekers frequently require care from mainstream health services demonstrating the importance of ensuring all sexual and reproductive health services are gender inclusive. Providers need training in specific needs, vulnerabilities, and preferences of the LGBTIQA+ community to enable patient-centred care. This involves providers confidently, sensitively, and pre-emptively consulting on use of preferred pronouns, sexual history, sexual orientation, and reproductive desires before engaging in care (Bartelt, 2020; Logie et al., 2018; Lowik, 2017).

The role of language when providing abortion care to LGBTIQA+ should not be underestimated. Organisations and providers need to understand client preferences before gendering body parts, referring to sexual partners as boyfriend or husband, and consider alternatives to labelling clinics as “women’s health”, which supports accessibility and safety (Bartelt, 2020). Additionally, gender inclusive in-take forms that move beyond gender binaries are crucial (Logie et al., 2018; Lowik, 2017).

Beyond abortion care, LGBTIQA+ people have inequitably poor access to contraception and are less likely to use contraception compared to cis-gendered people (Fix et al. 2020), likely contributing to abortion seeking. Failing to recognise a patient’s capacity to become pregnant can lead to an inaccurate assessment about risk of unintended pregnancy poor provision of care (Gomez et al., 2020; Ingraham, Wingo & Roberts, 2018).

More broadly, addressing the deep mistrust LGBTIQA+ people have developed because of past experiences with healthcare will require cross-sectoral commitment and resourcing. There is a need to acknowledge the adversity they have faced, the resilience they have drawn upon, and the negative outcomes they have endured (Mooney, 2017).

Our search did not result in any studies with a focus on pregnancy options and post-abortion counselling for LGBTIQA+ people, demonstrating a significant need for research in this space. Non-directive pregnancy options counselling services support pregnant people to make decisions about their pregnancy options with unbiased professional support. These services also support people post-abortion if required. Services in Australia identify that such counselling is often used by people experiencing complex socio-economic conditions or living with violence, reproductive coercion, or abuse (Children by Choice, 2020). Such services have been described by LGBTIQA+ participants as ‘life saving’ (Fix et al., 2020). Increasing accessibility to inclusive, non-directive pregnancy and abortion-related counselling services may be of critical importance to supporting the wellbeing of LGBTIQA+ people. Pregnancy decision making counselling services would benefit from greater research into the specific experiences and needs of LTBTIQA+ people when making pregnancy outcomes decisions and seeking support.

### Limitations

To minimise the risk of missed relevant articles, a scoping review methodology was employed, and a research librarian consulted in the development and application of the search strategy. A broader review including papers on inclusivity and safety within broader sexual and reproductive health services could lead to valuable insights into the experiences and needs of LGBTIQA+ individuals when accessing such services. In the current review, only papers published in English were included, which may have meant the exclusion of relevant studies published in other languages and may have contributed to the predominance of research from the USA.

## Conclusions

Access to safe and inclusive abortion care is a right of all, yet this review suggests it is a right not yet realised by many LGBTIQA+ people, even in contexts where abortion has been decriminalised. The LGBTIQA+ population face significant barriers to abortion, including systemic and individual level stigma and discrimination. The consequences of decades of poor care are many, but particularly alarming are the relatively high rates of unsafe self-induced abortion attempts reported, as the result of widespread healthcare avoidance and mistrust.

Along with challenges posited by health care systems design and service delivery, this review elucidated significant gaps in the extant literature, calling for a significant drive to conduct locally specific research to support a better understanding the needs of LGBTIQA+ people when accessing abortion care, pregnancy options and post-abortion counselling. There is a dearth of research exploring the specificities of need among LGBTIQA+ clients of pregnancy-options and post-abortion counselling service, which is vital to ensuring services are safe and accessible to this marginalised community. Alongside research and service-level changes, structural shifts are needed to combat discrimination in funding and health system design to support more holistic healthcare provision and outcomes for LGBTIQA+ people.

## Data Availability

All data produced in the present work are contained in the manuscript.

https://www.ncbi.nlm.nih.gov/pmc/articles/PMC7586656/

https://www.unboundmedicine.com/medline/citation/34238669/%22The_Health_System_Just_Wasn%27t_Built_for_Us%22:_Queer_Cisgender_Women_and_Gender_Expansive_Individuals%27_Strategies_for_Navigating_Reproductive_Health_Care_

https://pubmed.ncbi.nlm.nih.gov/32385584/

https://srh.bmj.com/content/48/e1/e22

https://www.sciencedirect.com/science/article/pii/S0002937820311261

https://pubmed.ncbi.nlm.nih.gov/28861537/

https://pubmed.ncbi.nlm.nih.gov/29661698/

